# Diagnostic testing and the evolution of detection avoidance by pathogens

**DOI:** 10.1101/2023.09.14.23295480

**Authors:** Jason Wood, Ben Ashby

## Abstract

Diagnostic testing is a key tool in the fight against many infectious diseases. The emergence of pathogen variants that are able to avoid detection by diagnostic testing therefore represents a key challenge for public health. In recent years, variants for multiple pathogens have emerged which escape diagnostic testing, including mutations in *Plasmodium falciparum* (malaria), *Chlamydia trachomatis* (chlamydia) and SARS-CoV-2 (COVID-19). However, little is currently known about when and the extent to which diagnostic test escape will evolve. Here we use a mathematical model to explore how the frequency of diagnostic testing, combined with variation in compliance and efficacy of quarantining, together drive the evolution of detection avoidance. We derive key thresholds under which a testing regime will (i) select for diagnostic test avoidance, or (ii) drive the pathogen extinct. Crucially, we show that imperfect compliance with diagnostic testing regimes can have marked effects on selection for detection avoidance, and consequently, for disease control. Yet somewhat counterintuitively, we find that an intermediate level of testing can select for the highest level of detection avoidance. Our results, combined with evidence from various pathogens, demonstrate that the evolution of diagnostic testing avoidance should be carefully considered when designing diagnostic testing regimes.

## Introduction

Pathogens may evolve in response to human interventions, which presents a key challenge for public health policymakers. For example, pharmaceutical interventions such as antimicrobials or vaccines, can lead to the evolution of antimicrobial resistance (AMR) (Davies and Davies 2010) or vaccine escape (Restif and Grenfell 2007), respectively. Pathogen evolution can therefore have dramatic effects on the efficacy of public health interventions by hindering efforts to treat or prevent infections. Consequently, substantial efforts have been devoted to understanding how pathogens evolve in response to public health policies, especially pharmaceutical interventions, and devising optimal strategies to mitigate or constrain pathogen evolution (Omenn 2010).

While much attention has been paid to the impact of pharmaceutical interventions on pathogen evolution, non-pharmaceutical interventions, such as diagnostic testing (Seth-Smith et al. 2009; Feleke et al. 2021) or reducing social mixing (Ashby, Smith, and Thompson 2023) may can also affect pathogen evolution. For example, routine surveillance of *Clostridium Difficile* in Costa Rica identified three variants which gave negative PCR test results, caused by deletion of the *tcdC* gene (Ramírez-Vargas et al. 2018). Similarly, in SARS-Cov-2, a deletion within the N gene can produce false negative antigen tests (Zannoli et al. 2022; Del Vecchio et al. 2022), and in *Plasmodium falciparum*, variants lacking the histidine-rich protein 2 (pfhrp2) and 3 (pfhrp3) genes have been identified following the introduction of rapid diagnostic tests targeting these antigens (Feleke et al. 2021). Similar patterns have been observed in sexually transmitted infections. In *Chlamydia trachomatis*, a variant with a 377 bp deletion has previously escaped diagnostic PCR testing (Seth-Smith et al. 2009; Persson et al. 2012), and in *Neisseria gonorrhoeae* it has been shown that diagnostic escape could occur through a reversion in the *gyrA* allele (the gene encoding the A subunit of DNA gyrase) (Rubin, Mortimer, and Grad 2023). Pathogen evolution arising from non-pharmaceutical interventions therefore has the potential for significant adverse effects on public health responses for many pathogens, akin to the effects of pharmaceutical interventions. Detection avoidance arising from diagnostic testing is of particular concern. Without reliable diagnostics, patients may experience prolonged symptoms or unknowingly transmit the disease. Understanding precisely how diagnostic testing affects pathogen evolution is crucial for assessing the risk of inducing selection for detection avoidance, and its impact on population health.

Theoretical studies on the evolution of detection avoidance have primarily focused on selection for asymptomatic infections (Saad-Roy et al. 2020; Okamoto et al. 2023), and by comparison relatively little is known about escape from diagnostic testing. As far as we are aware, only three studies have explored the evolution of detection avoidance arising from diagnostic testing. Watson et al. (2017) explored a mathematical model for the spread of a variant of *P. Falciparum* (the causative agent of malaria) which has been observed to escape Rapid Diagnostic Test kits (RDTs) due to gene deletions in histidine-rich protein 2 (HRP2). They found that selection for detection avoidance was most prevalent when malaria transmission was low, and patients were seeking and receiving treatment frequently based on diagnosis with RDTs. Smid et al. (2010) used a spatially-structured mathematical model of chlamydia transmission in Sweden to explore the emergence of a *C. trachomatis* variant known to escape detection by two diagnostic tests due to a plasmid deletion. They found that the *C. trachomatis* variant spread more favourably in counties which used both nucleic acid amplification tests, but once the testing regime had been updated to detect the new variant, the proportion of the population affected by the variant dramatically decreased. This could indicate a likely evolutionary trade-off associated with detection avoidance, which has also been speculated for the deletion of HRP2 in *P. falciparum* (Watson et al. 2017). Similarly, Del Vecchio et al. (2022) explored antigen test target failure in primary care settings in Italy for SARS-Cov-2. They found that a variant which produced false negative antigen tests was selected for in the region of Veneto, in which a higher proportion of tests were antigen rather than PCR. Del Vecchio et al. (2022) similarly found that the variant which produced false negative tests likely had a lower reproduction number than the Alpha variant, which was dominant at the time.

Here, we investigate how different testing regimes and the frequency or probability of taking a test affects selection for diagnostic test escape. In particular, we focus on how the effectiveness of (and compliance with) subsequent public health measures to reduce transmission (i.e., quarantine) mediates detection avoidance evolution. We derive key thresholds for (i) pathogen extinction due to testing, and (ii) detection avoidance to evolve, and show how evolution can prevent extinction. Crucially, we also show that an intermediate rate of testing can lead to the highest level of detection avoidance.

## Methods

### Model Description

We consider the epidemiological and evolutionary dynamics of a directly-transmitted pathogen in a well-mixed population of hosts, where *S* is the density of healthy (uninfected) hosts, *Q* and *A* are the densities of hosts who are aware of their infection and are either quarantining or not quarantining, respectively, *U* is the density of hosts who are not aware that they are infected, and *R* is the density of hosts who have recovered. The total density of the population is then *N* = *S*+ *Q* + *A* + *U*+ *R*. Hosts reproduce at a baseline rate *b*, which is not affected by infection status, but is subject to a density dependent crowding affect, *qN*, where *q* >0. Pathogen transmission is density-dependent at baseline rate *β*, with force of infection, *λ*(*ρ*) = *β*(*δQ*+ *A* +*U*), where *δ ∈* [0,1] is the reduction in transmission due to quarantine. We assume that there is no link between awareness of the infection and the rate of recovery, *γ*, or disease-associated mortality (virulence), α, and that hosts who are not aware of their infection do not change their behaviour. A full description of the model parameters can be found in Table 1.

We consider three versions of the model to account for different testing scenarios. In both cases, *ρ* is the probability of a false-negative test, which we assume to be under selection in the pathogen (see below), and *η* is the probability that an individual complies with quarantining measures. In the first version (Model A), all infected hosts take a single test with probability *σ∈*[0,1] when they initially become symptomatic, with the onset of symptoms coinciding with the beginning of the infectious period for all hosts. Upon infection, individuals with a positive test move into the quarantining (*Q*) class with probability *σ η*(1− *ρ*) and into the aware-but-not-quarantining class (*A*) with probability*σ*(1−*η*)(1− *ρ*)Individuals with a false-negative test move into the unaware class (*U*) with probability1− *σ*(1−*ρ*).The ecological dynamics for Model A are given by:

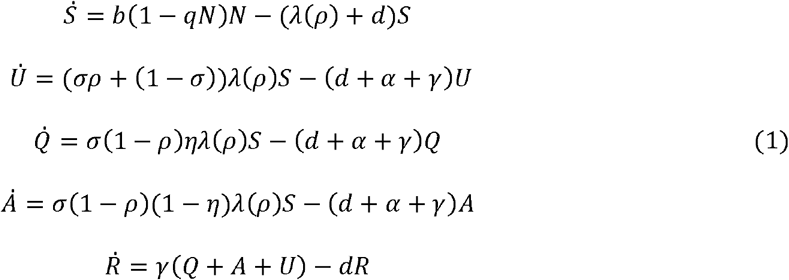

In the second version (Model B), infected hosts initially enter the unaware class (*U*) and test regularly at rate *ζ* with a probability*ρ* of returning a false negative per test (the outcome of tests are therefore independent). Infected individuals move to the quarantining class at rate *ζ*(1− *ρ*) *η* and to the aware-but-not-quarantining class (*A*) at rate *ζ*(1− *ρ*)(1− *η*). The ecological dynamics for Model B are given by:

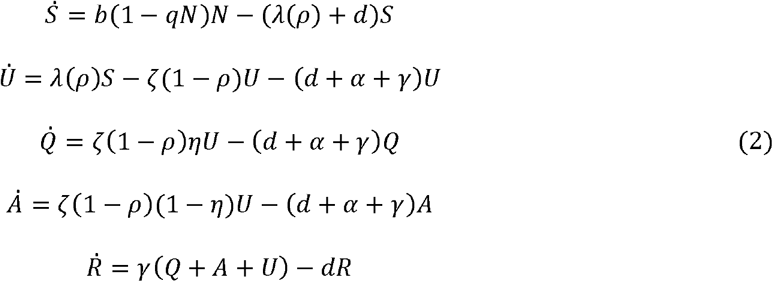

In the third version (Model C), hosts either enter a permanently unaware class (*U*) with probability(*ρ*) or the temporarily unaware class(*I*) with probability(1− *ρ*) Temporarily unaware hosts then move into the quarantining class (*Q*) at a rate *ζ η* and into the aware-but-not-quarantining class (*A*) with rate *ζ*(1−*η*). As the testing rate gets very large (*ζ*→∞), Model C becomes equivalent to Model A. In Model C, *N* = *S*+ *U*+ *I* + *A* + *Q* + *R* and *λ*(*ρ*) = *β*(*ρ*)(*δ Q* + *A* + *U*+ *I)*. The ecological dynamics of Model C are given by:

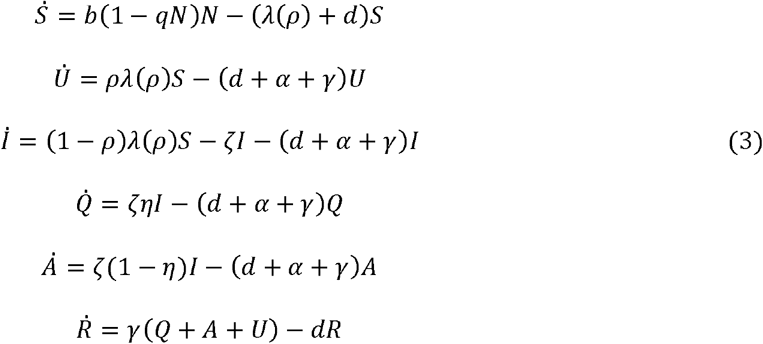

For all versions of the model, we investigate how compliance (*η*) and the effectiveness of quarantining (*δ*) affect selection for detection avoidance(*ρ*) in the pathogen. We assume there is a trade-off between pathogen transmission, *β*, and the likelihood of producing a false negative test, *ρ*, such that *β* = *β*(*ρ*) and hence *λ* = *λ*(*ρ*). Mechanistically, we are assuming that the more transmission stages that are produced, the more likely these are to be detected by the test, and so an increase in transmission comes at the cost of being more detectable, while detection-avoidance comes at the cost of a lower transmission rate, i.e.,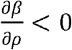. This assumption is motivated by data from chlamydia, SARS-Cov-2 and malaria infections, where variants that are less detectable by PCR and rapid testing also appear to be less transmissible (Persson et al. 2012; Feleke et al. 2021). There is no a priori reasoning for the curvature of the trade-off, so we explore both accelerating and decelerating trade-offs of the form,

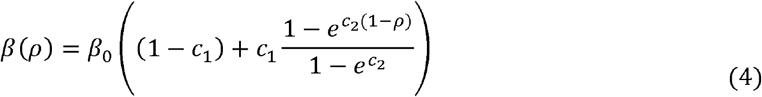

where *c*_1_ controls the strength of the trade-off and *c*_2_≠ 0 the curvature, with *c*_2_>0 corresponding to accelerating costs of detection avoidance on the transmission rate (diminishing returns of detection avoidance) and *c*_2_ > 0 corresponding to decelerating costs (increasing returns of detection avoidance).

**Table 1:** Parameter/ variable descriptions and default values.

| Parameter/<br>variable | Description | Default value(s) |
| --- | --- | --- |
| $b$ | Host birth rate | 2.0 |
| $c_1$ | Strength of trade-off | 1.0 |
| $c_2$ | Curvature of trade-off | -3.0 |
| $d$ | Host natural death rate | 0.1 |
| $m$ | Probability of new infection being a mutant | 0.2 |
| $q$ | Strength of density-dependent competition | 0.1 |
| $\alpha$ | Disease-associated mortality rate (virulence) | 0.5 |
| $\beta_0$ | Maximum pathogen transmission rate | 1.0 |
| $\gamma$ | Host recovery rate | 0.5 |
| $\delta$ | Effectiveness of quarantining measures | 0,0.1 |
| $\eta$ | Probability of host compliance with quarantining measures | 1,0.9 |
| $\lambda$ | Force of infection | n/a |
| $\rho$ | Probability of a false negative test | n/a |
| $\sigma$ | Probability of taking a test in Model A | 0.75 |
| $\zeta$ | Population testing rate in Models B and C | 3 |

We assume that mutations are sufficiently rare that the epidemiological dynamics (equations 1-3) reach a stable endemic equilibrium before a mutant arises, and that mutations have small phenotypic effects. A new mutant pathogen with false negativity probability *ρ*_*m*_ will then have dynamics,

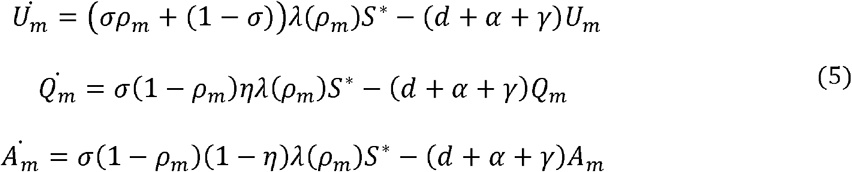

in Model A, and

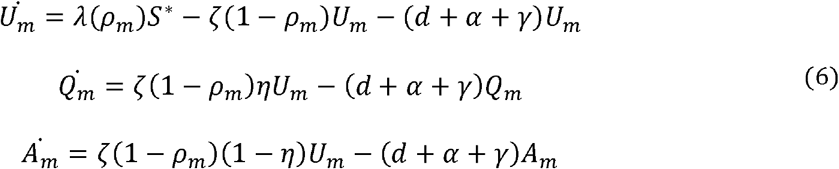

in Model B, and

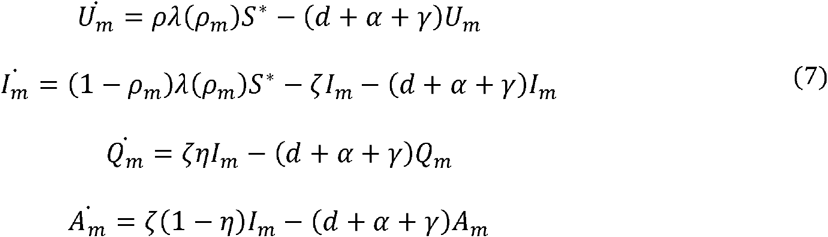

in Model C, where asterisks denote the resident population at equilibrium.

We calculate the invasion fitness, *r*(*ρ*_*m*_, *ρ*) of a rare mutant using the Next Generation Method (Hurford, Cownden, and Day 2010), and subsequently the fitness gradient, 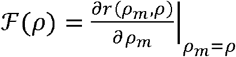 (see *Supplementary Material*), as:

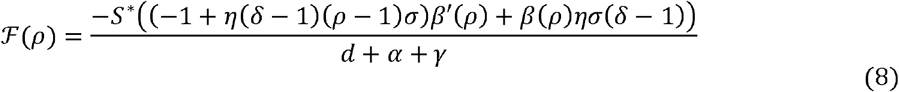

in Model A,

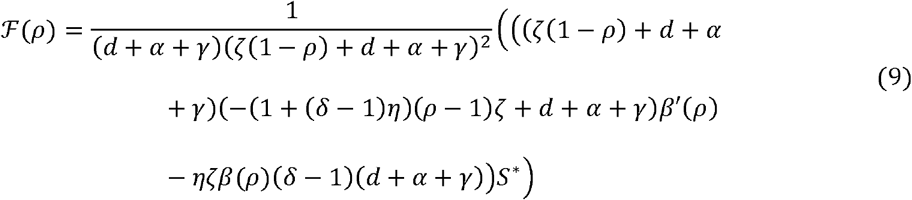

in Model B, and

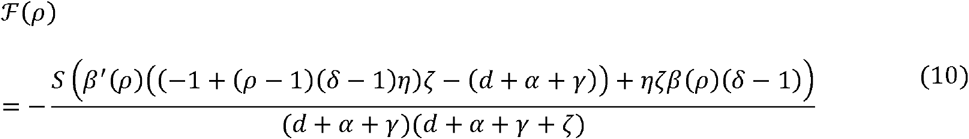

in Model C.

A singular strategy, *ρ*\*, is a value of which satisfies ℱ (*ρ*\*)= 0. The evolutionary stability of a singular strategy is determined by the sign of 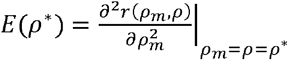, and the convergence stability is determined by the sign of *E*(*ρ*\*)+*M*(*ρ*\*) where 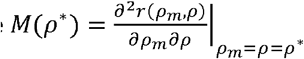. A singular strategy, *ρ*\*, is evolutionary stable if *E*(*ρ*\*)<0, and convergence stable if *E*(*ρ*\*)+*M*(*ρ*\*)< 0. In all three models it can be shown that 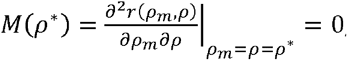, hence meaning any evolutionarily stable point is also convergence stable, and branching is not possible (see *Supplementary Material*).

Adaptive dynamics focuses on long-term evolutionary dynamics. To explore short-term evolutionary dynamics during the expansion phase of an epidemic, we used stochastic simulations by implementing the Gillespie algorithm (Gillespie 1977). Specifically, we create a discretised trait space for *ρ*, and assume that each new infection has a probability (*m*=0.2). of mutating to the adjacent strain. We assume the population size is large but non-replenishing (*N* = 100,000). We initialise simulations with 10 infected individuals and *N* − 10 susceptible individuals. We normalise each simulation by the time taken to reach the epidemic peak to compare across testing rates, as the time taken to reach the epidemic peak increases with the testing rate. At each recording step we calculate the average level of detection avoidance and track the number of infected hosts. For each set of parameter values we run twenty simulations. If the pathogen dies out before reaching at least 1,000 infected individuals at any given time, the simulation is discarded. We also compare our adaptive dynamics results to long-term stochastic simulations in a smaller, but replenishing population where any host that dies is replaced by a new susceptible, and recovered hosts re-enter the susceptible pool (*N* = 1000) . All code used to produce the figures within this paper is available within the Supplementary material and on GitHub.

## Results

### No compliance

Trivially, if there is no compliance with quarantining measures (*η* = 0), then the fitness gradient in all three models is

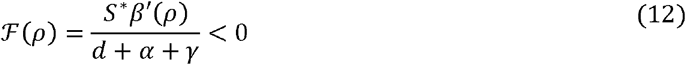

since *β*′(*ρ*) is negative for all *ρ*. The pathogen will therefore evolve to produce as few false negative tests as possible (*ρ* →0), favouring a higher rate of transmission (the same outcome occurs if quarantining has no effect on transmission (*δ*= 1)).

### Perfect compliance and quarantining

From the fitness gradients (equations 8-10), we can deduce that the strongest selection for false negative tests (highest value of *ρ*\*) will occur when there is perfect compliance with quarantining measures (*η*=1) and quarantining completely prevents transmission (*δ* = 1). In Model A, the singular strategy satisfies,

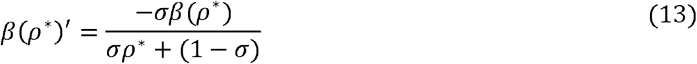

in Model B satisfies

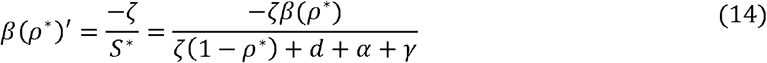

and in Model C satisfies

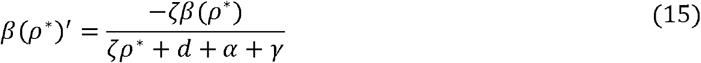

In Model A, there is only a single opportunity to move into the “aware” class as only one test is taken at the onset of symptoms, with probability *σ*. The probability of taking a test therefore plays a crucial role in driving selection for an intermediate probability of false positives, depending on the strength of the trade-off with transmission. In Model B, hosts test at a continuous rate leading to an additional flux out of the infectious class, similar to death (*d*+*α*) and recovery (*γ*). In Model C there is one opportunity to move into the aware classes, but the rate of testing varies. As a result, the optimal probability of false negativity depends on how the rate of testing *ζ* compares to the rate of movement out of the class due to other processes. When testing is maximal (*σ* = 1) in Model A and *ζ* → ∞ in Model B and Model C) any non-zero probability of false negative tests (*ρ* > 0) results in some individuals moving into the unaware infected class (*U*) in Model A and Model C, whereas in Model B the unaware class is only non-empty if the pathogen produces exclusively false negative tests (*ρ* = 1).

**Figure 1.**
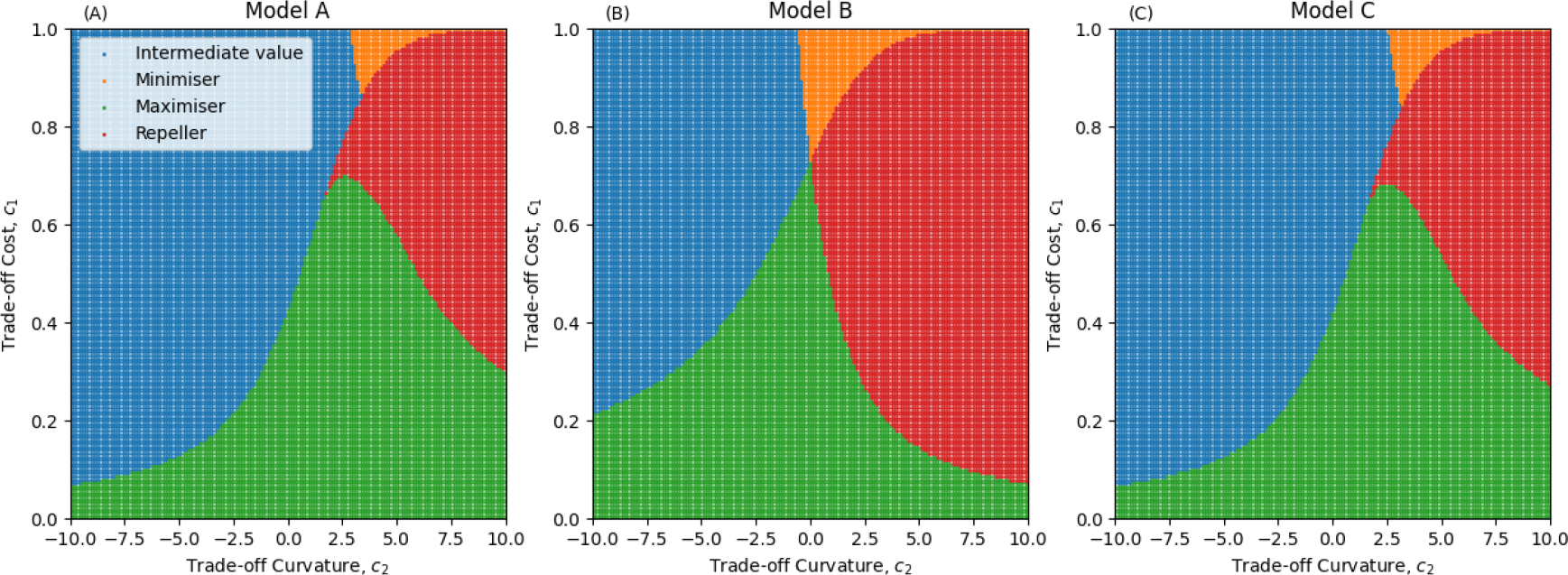
Evolutionary outcomes for Model A (A), Model B (B) and Model C (C), for different trade-off curvatures . and trade-off costs (c_1_).The pathogen experiences accelerating costs (diminishing returns) of detection avoidance when c_2_<0, and decelerating costs (increasing returns) when c_2_>0. Blue regions indicate the pathogen evolves to an intermediate level of detection avoidance (0 < ρ<1) The orange regions indicate where the pathogen does not evolve detection avoidance (ρ=0). In the green regions, the pathogen maximises detection avoidance (ρ = 1). In the red regions, there is an evolutionary repeller at an intermediate value of ρ, above which selection maximises detection avoidance (ρ=1) and below which selection minimises detection avoidance (ρ=0). Parameters as in Table 1 except where specified, δ = 0, η = 1

If detection avoidance is increasingly costly to transmission (*c*_2_<0), the pathogen always evolves to an evolutionarily stable strategy, including the boundaries *ρ*=0 and *ρ*=1 (Fig. 1). Accelerating costs imply diminishing returns, which generally favours an intermediate optimal level of detection avoidance. However, if detection avoidance is decreasingly costly (*c*_2_>0) the pathogen may evolve to either maximise (*ρ*=1) or minimise (*ρ*=0) detection avoidance depending on the initial conditions due to the presence of an evolutionary repeller. This is because decelerating costs mean there is a disproportionately high cost of false negativity when *ρ* is relatively small, which tends to select against detection avoidance, whereas there is a disproportionately low cost when *ρ* is sufficiently high, which tends to select for greater detection avoidance. Hereafter we will assume detection avoidance is increasingly costly (*c*_2_<0) as this is the only scenario where an intermediate level of false negativity may evolve in both models.

As we increase testing, we should always expect the optimal level of false negative tests to increase (in all of the models). However, if we imagine low testing, e.g., *σ*≈0 or *ζ*≈0, we would not expect to see selection for detection avoidance as there is a cost to transmission of producing false negative tests. From this we can infer that there is a threshold wherein testing needs to be sufficiently common before selection favours detection avoidance. In the *Supplementary Material* we derive the minimum test probability (Model A) and minimum testing rate (Models B and C) needed for detection avoidance to evolve. Similarly, if testing is sufficiently common (large *σ* or *ζ*), quarantining is sufficiently effective (large *δ*) and compliance is sufficiently high (large *η*), it may be possible for the pathogen to be driven extinct.

Unsurprisingly, more testing (between the minimum and extinction thresholds) selects for greater detection avoidance (Fig. 2A-C). Testing also reduces pathogen prevalence (Fig. 2D-F) and the basic reproduction number *R*_*0*_ (Fig. 2G-I), but evolution of detection avoidance allows the pathogen to persist at much higher levels of testing. Crucially, from a public health perspective, even if the pathogen evolves detection avoidance, testing still reduces pathogen prevalence. This occurs for two reasons. First, unless detection avoidance is perfect (*ρ*=1) or quarantine measures are completely ineffective (*η*=0 or *δ*=0), there is a non-zero probability that a test will be positive and result in reduced transmission due to quarantine. Second, detection avoidance is costly, resulting in an intrinsic reduction in transmission.

**Figure 2.**
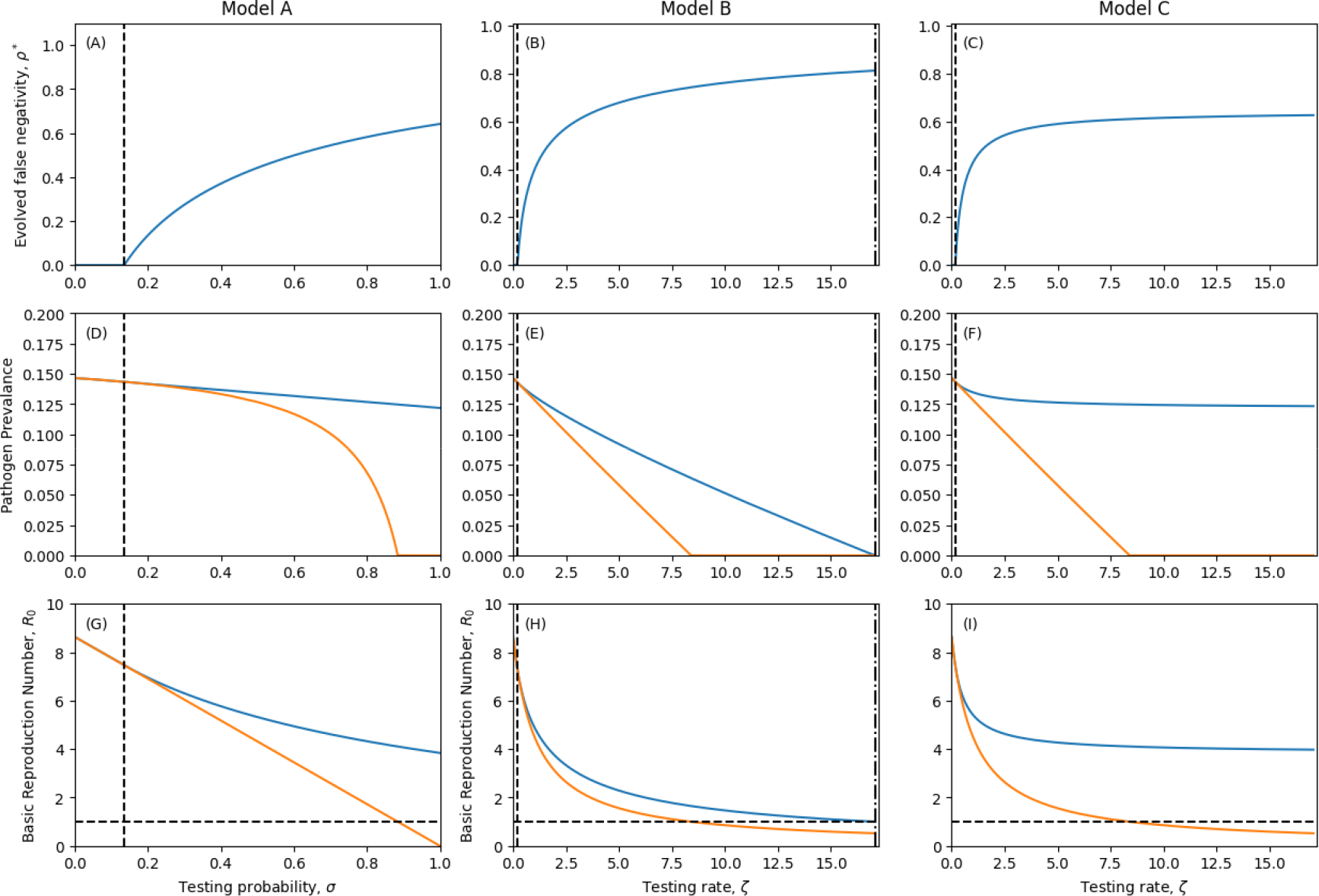
Evolved levels of false negativity under perfect compliance and quarantining (ρ* A-C), corresponding pathogen prevalence (proportion of hosts infected; D-F) and basic reproduction numbers (R_0_; G-I). The dashed vertical lines denote the level of testing required to select for detection avoidance (ρ > 0). Dot-dashed lines indicate pathogen extinction thresholds for the evolved pathogen (R_0_ < 1). In G-I, the horizontal dashed line indicates the extinction threshold R_0_ = 1. In D-I, the blue curve corresponds to the evolved pathogen and the orange curve to the ancestral state with no detection avoidance (ρ = 0). Parameters as in Table 1 except where specified δ = 0,η = 1.

### Imperfect quarantine

In reality, the effectiveness of quarantining and compliance with public health measures is imperfect (*δ*> 0, *η*< 1). Naturally, a decrease in compliance or the effectiveness of quarantining will shift the thresholds for extinction and the evolution of detection avoidance to higher testing probabilities (Model A) or rates (Models B and C). The testing threshold for the evolution of detection avoidance in Model A is closely related to the perfect compliance/quarantining scenario, with

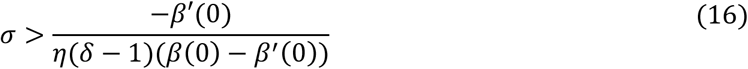

for detection avoidance to evolve. Clearly, when quarantine is perfect (*η*=1 and *δ*=0), this is equivalent to equation S53. Note that for sufficiently ineffective public health measures (whether in terms of compliance or the strictness of quarantine measures), the right-hand side of this inequality is greater than one and so there is no level of testing which selects for detection avoidance. The loss of perfect compliance and quarantining has no qualitative effect on the evolution of detection avoidance in Model A (Fig. 2A, Fig 3A) and in Model C, as the testing rate threshold is closely related to the perfect compliance / quarantining scenario with

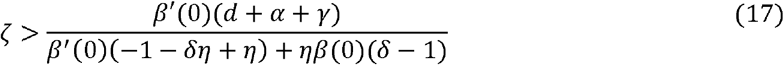

for detection avoidance to evolve. As in Model A, when quarantine and compliance is perfect, we recover equation S67.

In Model B, detection avoidance evolves provided

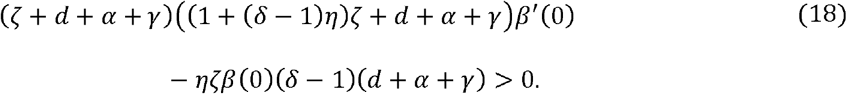

This inequality is quadratic in the testing rate, *ζ*, and so can be solved to find upper and lower bounds for when testing selects for detection avoidance. Detection avoidance therefore peaks for an intermediate testing rate in Model B (Fig. 3B), in contrast to the perfect compliance and quarantining scenario (Fig. 2B).

This result is somewhat unintuitive, as one might expect that an increase in testing always selects for greater detection avoidance (as in Fig. 2B). However, this does not account for the costs associated with false negative tests and the “leakiness” that occurs with imperfect quarantining. As the testing rate becomes sufficiently large, infected individuals rapidly leave the unaware class. Hence, production of new infections from the aware classes (either due to imperfect quarantining or some hosts not quarantining) becomes increasingly important, with detection avoidance less advantageous due to the independence between tests.

This can be thought of as a shifting of priorities. When testing is rare, selection favours high transmissibility and low detection avoidance as infected individuals spend most of their time in the unaware class (detection avoidance is not necessary). As testing becomes more common, selection favours detection avoidance as this increases time spent in the unaware class where onwards transmission is greatest. But as testing becomes even more frequent, selection once again favours higher transmission and lower detection avoidance as most infected individuals quickly move into the aware classes even if the pathogen produces false negative tests, because the result of each test is independent. If an infected individual takes *n* tests, then the probability that all tests are negative is *ρ*^*n*^ which tends to 0 as *n*→∞. provided *ρ*≠1, and hence a higher testing rate renders detection avoidance ineffective if the results of each test are independent.

In contrast to perfect quarantining and compliance (Fig. 2), imperfect quarantining may be insufficient to drive the pathogen extinct, even if testing occurs at a very high rate and there is no detection avoidance (Fig. 3D-F). This is because the pathogen may be completely sustained by infections produced in the aware classes. Our observations indicate that for even slightly imperfect quarantining or compliance, the testing rate needed to cause extinction is unreasonably high. We also note that while testing always reduces the prevalence of the pathogen, the imperfect nature of quarantining and compliance with public health measures may mean that the prevalence does not drastically reduce (Fig. 3D-F). This indicates that the effectiveness of diagnostic tests for reducing disease prevalence may be limited.

**Figure 3.**
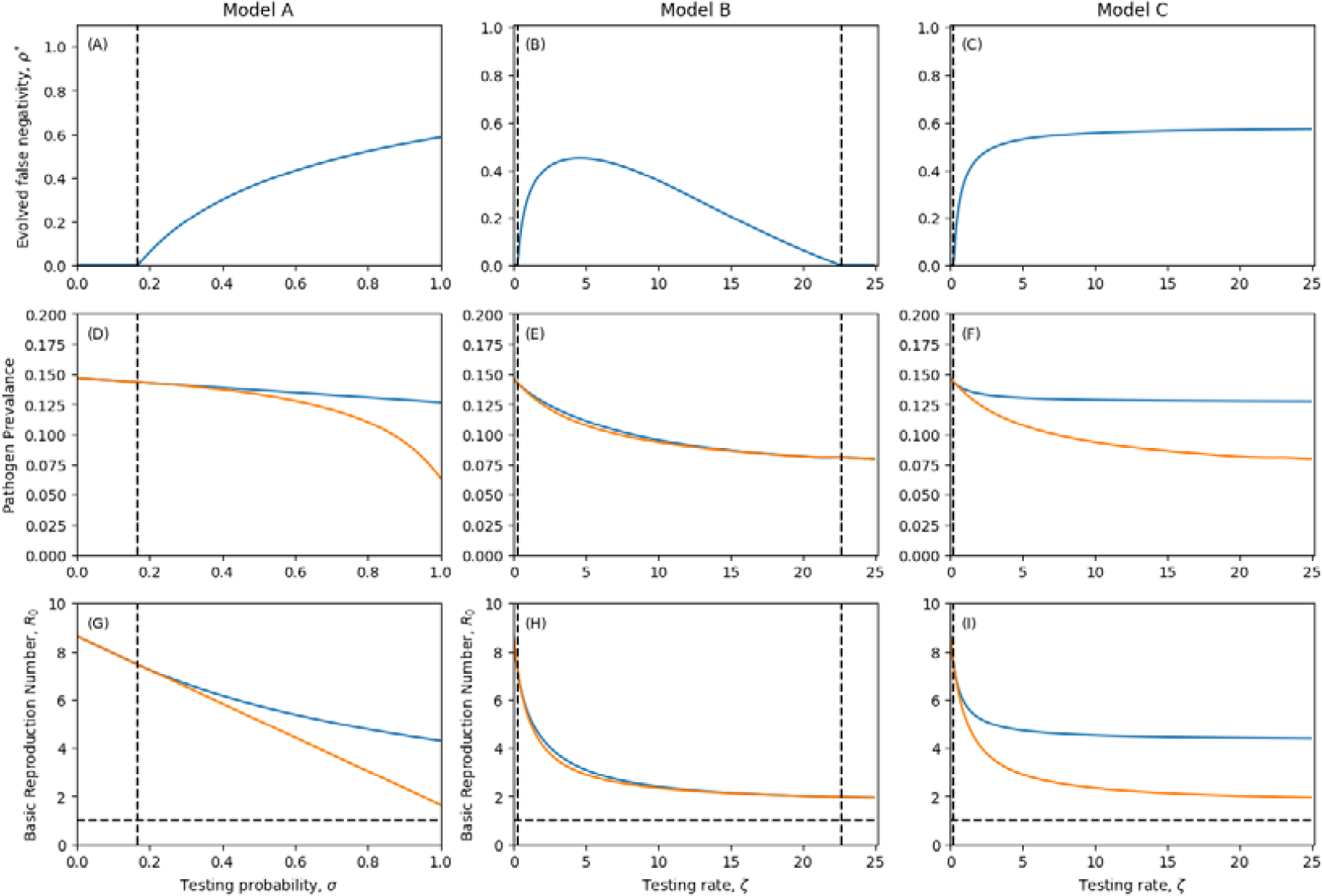
Evolved levels of false negativity with imperfect quarantining (A-C), corresponding pathogen prevalence (D-F) and reproduction numbers (G-I). Dashed vertical line denotes level of testing required to select for false negative tests. Dotted and Dashed line indicates extinction thresholds. In Reproduction number plots a horizontal dashed line indicates the value R_0_ = 1. In pathogen prevalence and reproduction number plot, blue line indicates prevalence or R_0_ of singular strategy, orange line indicates prevalence or R_0_ in the absence of evolution. Parameters as in Table 1 except where specified, δ = 0.1, η = 0.9.

### Stochastic Simulations

Our long-term adaptive dynamics predictions are consistent in stochastic simulations in smaller, finite populations with replenishment, with detection avoidance increasing with greater testing in Models A and C but peaking at intermediate testing in Model B (Fig. S1). The short-term evolutionary dynamics in a large but non-replenishing population are broadly similar in Models A and C, with detection avoidance increasing with greater testing (Fig. 4). However, in Model A we do observe a substantial increase in detection avoidance when every individual is tested (*σ*=1), which does not occur in our adaptive dynamics analysis. This suggests that extreme levels of testing create very strong short-term selection for detection avoidance, but this recedes over the long term. In Model C we see no significant differences between our adaptive dynamics analysis and the results of stochastic simulations. In Model B, however, selection for detection avoidance saturates as the testing rate increases rather than peaking at intermediate testing rates (Fig. S2). This suggests that during the expansion phase of an epidemic, there may still be an advantage to detection avoidance even if hosts are frequently tested with each test having an independent result, as there is still a large pool of susceptible hosts that has yet to be sufficiently depleted.

**Figure 4.**
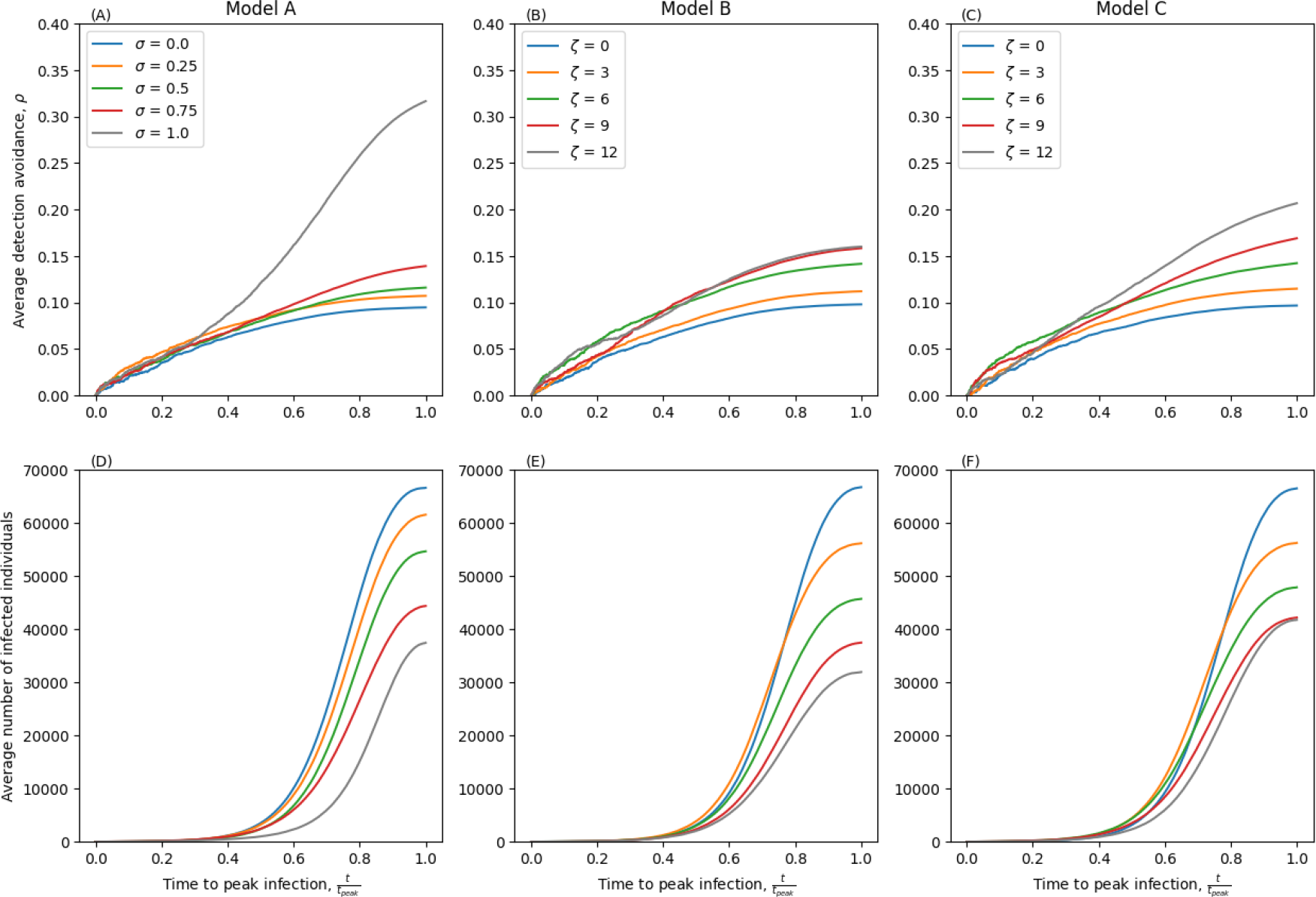
Stochastic simulations of Models A-C with imperfect quarantining in a large, non-replenishing population. The first row of figures (A-C) show the average detection avoidance at a given time. The second row of figures (D-F) show the average number of infected individuals. Parameters as in Table 1 except δ = 0.1, η = 1.0, β = 0.0001, N = 100,000.

## Discussion

The evolution of pathogens in response to human interventions is well documented (Restif and Grenfell 2007; Seth-Smith et al. 2009; Davies and Davies 2010; Persson et al. 2012; Feleke et al. 2021; Zannoli et al. 2022). Intuitively, efforts to manage infectious diseases naturally select for pathogen variants that can resist or evade interventions. The evolution of anti-microbial resistance (AMR) (Davies and Davies 2010) and vaccine escape (Restif and Grenfell 2007) are the most prominent examples. Yet pathogens can also evolve in response to non-pharmaceutical interventions (Ashby, Smith, and Thompson 2023). In particular, there is a growing body of evidence that many pathogens can readily evolve in response to diagnostic testing, but little is known about precisely how diagnostic testing affects selection for detection avoidance. As far as we are aware, only three theoretical studies (Seth-Smith et al. 2009; Watson et al. 2017; Del Vecchio et al. 2022) have considered the evolution of detection avoidance arising from diagnostic testing, and these have focused on specific pathogens rather than the development of general eco-evolutionary theory of detection avoidance.

Here we have explored how three approaches to diagnostic testing drive the evolution of detection avoidance. When infected individuals may only take a single test (Models A and C), there is a minimum threshold for selection for detection avoidance, above which more testing always results in selection for higher false negativity. In some cases, a sufficiently high level of testing may drive the pathogen extinct. In contrast, when infected individuals regularly test for infection but test results are independent (Model B), more frequent testing does not necessarily lead to selection for higher false negativity. When either quarantining or compliance are imperfect, detection avoidance peaks at an intermediate testing rate in Model B, with higher testing rates selecting for low levels of detection avoidance in the long-term. In the perfect quarantining and compliance scenario, an increase in the testing rate leads to more “dead-end” infections by fully preventing onwards transmission, and hence detection avoidance increases with the testing rate. But in the imperfect quarantining and compliance scenario, aware individuals can still produce new infections. Hence, as the testing rate increases, selection shifts from prioritising transmission before quarantining (investment in detection avoidance) to transmission in the aware class (investment in infectivity). However, our stochastic simulations reveal that the latter outcome only occurs in the long-term, as detection avoidance instead saturates as the testing rate increases in the short-term. This suggests that during the epidemic phase, selection for detection avoidance is higher than during the endemic phase, most likely due to there still being a large pool of susceptible hosts to exploit. This is analogous to virulence evolution, where move virulent strains may have a growth advantage during the early stages of an epidemic, but lose out in the long run when hosts become more scarce (see e.g., Berngruber et al. 2013).

Although the evolution of detection avoidance from diagnostic testing presents a significant public health concern, in our models testing still always reduces overall pathogen prevalence in the population relative to the absence of testing. This happens for two reasons. First, unless the pathogen completely avoids detection, testing inevitably leads to a reduction in transmission as some individuals will quarantine. Second, because we assumed that there is a trade-off between false negativity and transmissibility – motivated by evidence from pathogens including *P. falciparum* (Watson et al. 2017), *C. trachomatis* (Smid et al. 2020), and SARS-CoV-2 (Del Vecchio et al. 2022) – avoiding detection is costly. Detection avoidance therefore never results in an increase in pathogen prevalence relative to the absence of testing. However, the emergence of detection avoidance would clearly result in an increase in pathogen prevalence at a given level of testing.

Our model made a number of key assumptions which should be relaxed and explored in future work. Most importantly, when using an adaptive dynamics framework, which employs a separation of ecological and evolutionary timescales, we were not able to explore selection for detection avoidance during transient epidemic dynamics. However, this approach does provide qualitative insights into the effects of healthcare interventions on the direction of selection (see also Fan and Geritz (2023)), and the adaptive dynamics assumptions were relaxed in our stochastic simulations. We also assumed that the host population was homogeneous, but significant heterogeneity in both disease outcomes and compliance with NPIs is likely in most populations. For example, during the COVID-19 pandemic there was considerable variation in compliance with NPIs (Seale et al. 2020). Such heterogeneity may weaken the effects of diagnostic testing on transmission (similar to reducing *η* or *δ* in our model), and hence generally weaken selection for detection avoidance. The effects of awareness-driven behaviour changes at the population level can also lead to complex feedbacks on epidemic dynamics, but these effects have received relatively little attention (Weitz et al. 2020). Finally, a key assumption in Model B is that the outcome of each test is independent of all others. Hence, repeated testing at a high enough rate would eventually detect most (if not all) infections (provided *ρ*<1). This was based on the assumption that detection avoidance is probabilistic, but if tests always yield the same result for a given infection, then the peak in detection avoidance at intermediate testing rates does not occur, as observed in Model C. Finally, we did not consider the economic costs or side effects of testing, including the impact of social isolation due to quarantining, or the impact of false positive tests on the population, as our focus was on the evolutionary effects of diagnostic testing.

This study was motivated by evidence of detection avoidance arising from diagnostic testing in a variety of pathogens (Seth-Smith et al. 2009; Persson et al. 2012; Ramírez-Vargas et al. 2018; Feleke et al. 2021; Zannoli et al. 2022; Rubin, Mortimer, and Grad 2023). Rapid antigenic and PCR testing became particularly widespread during the COVID-19 pandemic, and this may signal a shift in attitudes towards (and the availability of) diagnostic tests for other pathogens. Interestingly, as new major variants emerged during the COVID-19 pandemic, a cyclical pattern emerged in spike gene target failures (SGTF) in PCR tests, which allowed for accurate identification of specific variants (McMillen et al. 2022). While this did not lead to detection avoidance, clearly the spike gene was under selection (although cycles in SGTF could have arisen due to interference competition; i.e., the Hill-Robertson effect). One might speculate that had the spike gene been the only target of PCR tests, variants with SGTF would have been at an even greater advantage due to detection avoidance. Conversely, the multi-pronged testing regime used in many countries during the COVID-19 pandemic (frequent rapid antigen tests and PCR testing with multiple target genes) is likely a good strategy to help mitigate the emergence of variants that can avoid diagnostic tests.

While theoretical studies of detection avoidance evolution are rare, there are strong analogies with sexually transmitted infections (STIs), such as chlamydia and gonorrhoea, which may have evolved to be inconspicuous to avoid detection by choosy mates (Ashby et al. 2019, Pirrie, Chapman, and Ashby 2022). If prospective mates are less likely to choose partners who display signs of infection, then this can create strong selection for being inconspicuous, similar to selection for diagnostic testing avoidance. For example, Syphilis, which initially presented as a severely acute and noticeable disease during the middle ages, rapidly evolved to be much more cryptic, likely due to mate-choice effects in humans (Knell 2004). Thus, the patterns we see in our model are in good agreement with established evolutionary theory.

Overall, we have shown how the nature of testing, compliance with public health measures to reduce transmission, and the efficacy of quarantining measures, can dramatically drive selection for detection avoidance. In particular, our results demonstrate that detection avoidance can readily evolve above a threshold level of testing, but also that selection for detection avoidance can weaken, or even change direction, at higher levels of testing. However, since diagnostic testing always reduces disease prevalence, it is likely to be beneficial despite the evolution of detection avoidance.

## Data Availability

All data produced in the present work can be recreated using the source code available at the associated github link

https://github.com/JasonRWood/evolution_of_detection_avoidance

## Acknowledgements

JW is supported by a scholarship from the EPSRC Centre for Doctoral Training in Statistical Applied Mathematics at Bath (SAMBa), under the project EP/L015684/1. BA is supported by the Natural Environment Research Council (grant numbers NE/N014979/1 and NE/V003909/1). We acknowledge the support of the Natural Sciences and Engineering Research Council of Canada (NSERC). Nous remercions le Conseil de recherches en sciences naturelles et en génie du Canada (CRSNG) de son soutien.

