## Supplementary_Material for "Diagnostic testing and the evolution of detection avoidance by pathogens"

---


**1 Derivation of fitness gradients ..... 2**

**1.1 Model A ..... 2**

**1.2 Model B ..... 3**

**1.3 Model C ..... 3**

**2 Stability analysis of singular strategies ..... 5**

**2.1 Model A ..... 5**

**2.2 Model B ..... 6**

**2.3 Model C ..... 7**

**3 Selection for detection avoidance ..... 8**

**3.1 Model A ..... 8**

3.1.1 Perfect compliance and quarantining ..... 8

3.1.2 Imperfect quarantining ..... 8

**3.2 Model B ..... 9**

3.2.1 Perfect compliance and quarantining ..... 9

3.2.2 Imperfect quarantining ..... 10

**3.3 Model C ..... 11**

3.3.1 Perfect compliance and quarantining ..... 11

3.3.2 Imperfect quarantining ..... 11

**4 Supplementary figures ..... 12**

### 1 Derivation of fitness gradients

#### *1.1 Model A*

The rare mutant dynamics for Model A are described by

$$\dot{U_m} = \big(\sigma\rho_m + (1-\sigma)\big)\lambda(\rho_m)S^* - (d+\alpha+\gamma)U_m \quad \text{S1}$$
$$\dot{A_m} = \sigma(1-\rho_m)(1-\eta)\lambda(\rho_m)S^* - (d+\alpha+\gamma)A_m \quad \text{S2}$$
$$\dot{Q_m} = \sigma(1-\rho_m)\eta\lambda(\rho_m)S^* - (d+\alpha+\gamma)Q_m \quad \text{S3}$$

with Jacobian

$$J = \begin{pmatrix} \big(\sigma\rho_m + (1-\sigma)\big)\beta(\rho_m)S^* - (d+\alpha+\gamma) & \big(\sigma\rho_m + (1-\sigma)\big)\beta(\rho_m)S^* & \big(\sigma\rho_m + (1-\sigma)\big)\delta\beta(\rho_m)S^* \\ \sigma(1-\rho_m)\eta\beta(\rho_m)S^* & \sigma(1-\rho_m)\eta\beta(\rho_m)S^* - (d+\alpha+\gamma) & \sigma(1-\rho_m)\eta\delta\beta(\rho_m)S^* \\ \sigma(1-\rho_m)(1-\eta)\beta(\rho_m)S^* & \sigma(1-\rho_m)(1-\eta)\beta(\rho_m)S^* & \sigma(1-\rho_m)(1-\eta)\delta\beta(\rho_m)S^* - (d+\alpha+\gamma) \end{pmatrix} \quad \text{S4}$$

From this we can derive the next generation matrix, $NG = FV^{-1}$, where:

$$F = \begin{pmatrix} \big(\sigma\rho_m + (1-\sigma)\big)\beta(\rho_m)S^* & \big(\sigma\rho_m + (1-\sigma)\big)\beta(\rho_m)S^* & \big(\sigma\rho_m + (1-\sigma)\big)\delta\beta(\rho_m)S^* \\ \sigma(1-\rho_m)\eta\beta(\rho_m)S^* & \sigma(1-\rho_m)\eta\beta(\rho_m)S^* & \sigma(1-\rho_m)\eta\delta\beta(\rho_m)S^* \\ \sigma(1-\rho_m)(1-\eta)\beta(\rho_m)S^* & \sigma(1-\rho_m)(1-\eta)\beta(\rho_m)S^* & \sigma(1-\rho_m)(1-\eta)\delta\beta(\rho_m)S^* \end{pmatrix} \quad \text{S5}$$

$$V = F - J = \begin{pmatrix} d+\alpha+\gamma & 0 & 0 \\ 0 & d+\alpha+\gamma & 0 \\ 0 & 0 & d+\alpha+\gamma \end{pmatrix} \quad \text{S6}$$

$$V^{-1} = \begin{pmatrix} \frac{1}{d+\alpha+\gamma} & 0 & 0 \\ 0 & \frac{1}{d+\alpha+\gamma} & 0 \\ 0 & 0 & \frac{1}{d+\alpha+\gamma} \end{pmatrix} \quad \text{S7}$$

$$NG = \begin{pmatrix} \frac{\big(\sigma\rho_m + (1-\sigma)\big)\beta(\rho_m)S^*}{d+\alpha+\gamma} & \frac{\big(\sigma\rho_m + (1-\sigma)\big)\beta(\rho_m)S^*}{d+\alpha+\gamma} & \frac{\big(\sigma\rho_m + (1-\sigma)\big)\delta\beta(\rho_m)S^*}{d+\alpha+\gamma} \\ \frac{\sigma(1-\rho_m)\eta\beta(\rho_m)S^*}{d+\alpha+\gamma} & \frac{\sigma(1-\rho_m)\eta\beta(\rho_m)S^*}{d+\alpha+\gamma} & \frac{\sigma(1-\rho_m)\eta\delta\beta(\rho_m)S^*}{d+\alpha+\gamma} \\ \frac{\sigma(1-\rho_m)(1-\eta)\beta(\rho_m)S^*}{d+\alpha+\gamma} & \frac{\sigma(1-\rho_m)(1-\eta)\beta(\rho_m)S^*}{d+\alpha+\gamma} & \frac{\sigma(1-\rho_m)(1-\eta)\delta\beta(\rho_m)S^*}{d+\alpha+\gamma} \end{pmatrix} \quad \text{S8}$$

The largest eigenvalue of the next generation matrix, and thus our sign equivalent expression for fitness is then,

$$r(\rho_m, \rho) = -\frac{\beta(\rho_m)S^*(\rho_m\eta\sigma\delta - \rho_m\eta\sigma - \eta\sigma\delta + \eta\sigma - 1)}{d+\alpha+\gamma} - 1 \quad \text{S9}$$

And hence the fitness gradient is,

$$\mathcal{F}(\rho) = \left.\frac{dr}{d\rho_m}\right|_{\rho_m=\rho} = \frac{-S^*\left((-1+\eta(\delta-1)(\rho-1)\sigma)\frac{d\beta(\rho)}{d\rho} + \beta(\rho)\eta\sigma(\delta-1)\right)}{d+\alpha+\gamma} \quad \text{S10}$$

#### 1.2 Model B

The rare mutant dynamics are described by

$$\dot{U_m} = \lambda(\rho_m)S^* - \zeta(1-\rho_m)U_m - (d+\alpha+\gamma)U_m \quad \text{S11}$$
$$\dot{A_m} = \zeta(1-\rho_m)(1-\eta)U_m - (d+\alpha+\gamma)A_m \quad \text{S12}$$
$$\dot{Q_m} = \zeta(1-\rho_m)\eta U_m - (d+\alpha+\gamma)Q_m \quad \text{S13}$$

with Jacobian

$$J = \begin{pmatrix} \beta(\rho_m)S^* - \zeta(1-\rho_m) - (d+\alpha+\gamma) & \beta(\rho_m)S^* & \delta\beta(\rho_m)S^* \\ \zeta(1-\rho_m)\eta & -(d+\alpha+\gamma) & 0 \\ \zeta(1-\rho_m)(1-\eta) & 0 & -(d+\alpha+\gamma) \end{pmatrix} \quad \text{S14}$$

Using the same method as above, we can calculate the next generation matrix, $NG = FV^{-1}$, where

$$F = \begin{pmatrix} \beta(\rho_m)S^* & \beta(\rho_m)S^* & \delta\beta(\rho_m)S^* \\ 0 & 0 & 0 \\ 0 & 0 & 0 \end{pmatrix} \quad \text{S15}$$

$$V = F - J = \begin{pmatrix} \zeta(1-\rho_m) + (d+\alpha+\gamma) & 0 & 0 \\ -\zeta(1-\rho_m)\eta & (d+\alpha+\gamma) & 0 \\ -\zeta(1-\rho_m)(1-\eta) & 0 & (d+\alpha+\gamma) \end{pmatrix} \quad \text{S16}$$

$$V^{-1} = \begin{pmatrix} \frac{-1}{\zeta(1-\rho_m) + (d+\alpha+\gamma)} & 0 & 0 \\ \frac{\zeta(1-\rho_m)\eta}{(d+\alpha+\gamma)\left(\zeta(1-\rho_m) + (d+\alpha+\gamma)\right)} & \frac{1}{d+\alpha+\gamma} & 0 \\ \frac{\zeta(1-\rho_m)(\eta-1)}{(d+\alpha+\gamma)\left(\zeta(1-\rho_m) + (d+\alpha+\gamma)\right)} & 0 & \frac{1}{d+\alpha+\gamma} \end{pmatrix} \quad \text{S17}$$

$$NG = \begin{pmatrix} \frac{\delta\beta(\rho_m)S^*\zeta(\rho_m-1)\eta}{(d+\alpha+\gamma)\left(\zeta(\rho_m-1) - (d+\alpha+\gamma)\right)} - \frac{\beta(\rho_m)S^*\zeta(\rho_m-1)(\eta-1)}{(d+\alpha+\gamma)\left(\zeta(\rho_m-1) - (d+\alpha+\gamma)\right)} - \frac{\beta(\rho_m)S^*}{\zeta(\rho_m-1) - (d+\alpha+\gamma)} & \frac{\beta(\rho_m)S^*}{d+\alpha+\gamma} & \frac{\delta\beta(\rho_m)S^*}{d+\alpha+\gamma} \\ 0 & 0 & 0 \\ 0 & 0 & 0 \end{pmatrix} \quad \text{S18}$$

From which we can derive an expression which is sign equivalent to fitness minus one

$$r(\rho_m, \rho) = \frac{\beta(\rho_m)S^*(\delta\zeta\eta\rho_m - \rho_m\zeta\eta - \delta\zeta\eta + \rho_m\zeta + \zeta\eta - (d+\alpha+\gamma) - \zeta)}{(d+\alpha+\gamma)\left(\zeta(\rho_m-1) - (d+\alpha+\gamma)\right)} - 1 \quad \text{S19}$$

and hence the fitness gradient is:

$$\mathcal{F}(\rho) = \frac{1}{(d+\alpha+\gamma)(\zeta(1-\rho)+d+\alpha+\gamma)^2}\Big(\big((\zeta(1-\rho)+d+\alpha +\gamma)(-(1+(\delta-1)\eta)(\rho-1)\zeta + d+\alpha+\gamma)\beta'(\rho) - \eta\zeta\beta(\rho)(\delta-1)(d+\alpha+\gamma)\big)S^*\Big) \quad \text{S20}$$

#### 1.3 Model C

The mutant dynamics are described by

$$\dot{U_m} = \rho_m\lambda(\rho_m)S^* - (d+\alpha+\gamma)U_m \quad \text{S21}$$
$$\dot{I_m} = (1-\rho_m)\lambda(\rho_m)S^* - \zeta I_m - (d+\alpha+\gamma)I_m \quad \text{S22}$$

$$\dot{A_m} = \zeta(1-\eta)I_m - (d+\alpha+\gamma)A_m \quad \text{S23}$$
$$\dot{Q_m} = \zeta\eta I_m - (d+\alpha+\gamma)Q_m \quad \text{S24}$$

with Jacobian

$$J = \begin{pmatrix} \rho_m\beta(\rho_m)S^* - (d+\alpha+\gamma) & \rho_m\beta(\rho_m)S^* & \rho\beta(\rho_m)S^* & \delta\rho_m\beta(\rho_m)S^* \\ (1-\rho_m)\beta(\rho_m)S^* & (1-\rho_m)\beta(\rho_m)S^* - \zeta - (d+\alpha+\gamma) & (1-\rho_m)\beta(\rho_m)S^* & (1-\rho_m)\delta\beta(\rho_m)S^* \\ 0 & \zeta(1-\eta) & -(d+\alpha+\gamma) & 0 \\ 0 & \zeta\eta & 0 & -(d+\alpha+\gamma) \end{pmatrix} \quad \text{S25}$$

From this we can derive the next generation matrix, $NG = FV^{-1}$, where:

$$F = \begin{pmatrix} \rho_m\beta(\rho_m)S^* & \rho_m\beta(\rho_m)S^* & \rho\beta(\rho_m)S^* & \delta\rho_m\beta(\rho_m)S^* \\ (1-\rho_m)\beta(\rho_m)S^* & (1-\rho_m)\beta(\rho_m)S^* & (1-\rho_m)\beta(\rho_m)S^* & (1-\rho_m)\delta\beta(\rho_m)S^* \\ 0 & 0 & 0 & 0 \\ 0 & 0 & 0 & 0 \end{pmatrix} \quad \text{S26}$$

$$V = F - J = \begin{pmatrix} d+\alpha+\gamma & 0 & 0 & 0 \\ 0 & \zeta+d+\alpha+\gamma & 0 & 0 \\ 0 & -\zeta(1-\eta) & d+\alpha+\gamma & 0 \\ 0 & -\zeta\eta & 0 & d+\alpha+\gamma \end{pmatrix} \quad \text{S27}$$

$$V^{-1} = \begin{pmatrix} \frac{1}{d+\alpha+\gamma} & 0 & 0 & 0 \\ 0 & \frac{1}{\zeta+d+\alpha+\gamma} & 0 & 0 \\ 0 & \frac{-\zeta(1-\eta)}{(\zeta+d+\alpha+\gamma)(d+\alpha+\gamma)} & \frac{1}{d+\alpha+\gamma} & 0 \\ 0 & \frac{\zeta\eta}{(\zeta+d+\alpha+\gamma)(d+\alpha+\gamma)} & 0 & \frac{1}{d+\alpha+\gamma} \end{pmatrix} \quad \text{S28}$$

$$NG = \begin{pmatrix} \frac{\rho_m\beta(\rho_m)S^*}{d+\alpha+\gamma} & \frac{\rho_m\beta(\rho_m)S^*((1+(\delta-1)\eta)\zeta+d+\alpha+\gamma)}{(d+\alpha+\gamma)(\zeta+d+\alpha+\gamma)} & \frac{\rho_m\beta(\rho_m)S^*}{d+\alpha+\gamma} & \frac{\delta\rho_m\beta(\rho_m)S^*}{d+\alpha+\gamma} \\ \frac{(1-\rho_m)\beta(\rho_m)S^*}{d+\alpha+\gamma} & \frac{(1-\rho_m)\beta(\rho_m)S^*((1+(\delta-1)\eta)\zeta+d+\alpha+\gamma)}{(d+\alpha+\gamma)(\zeta+d+\alpha+\gamma)} & \frac{(1-\rho_m)\beta(\rho_m)S^*}{d+\alpha+\gamma} & \frac{\delta(1-\rho_m)\beta(\rho_m)S^*}{d+\alpha+\gamma} \\ 0 & 0 & 0 & 0 \\ 0 & 0 & 0 & 0 \end{pmatrix} \quad \text{S29}$$

The largest eigenvalue of the next generation matrix, and thus our sign equivalent expression for fitness is then,

$$r(\rho_m, \rho) = -\frac{S^*\big((-1+(-1+\rho_m)(\delta-1)\eta)\zeta - (d+\alpha+\gamma)\big)\beta(\rho_m) - (d+\alpha+\gamma)(\zeta+d+\alpha+\gamma)}{(d+\alpha+\gamma)(\zeta+d+\alpha+\gamma)} \quad \text{S30}$$

And hence the fitness gradient is,

$$\mathcal{F}(\rho) = \frac{\big((-1+(-1+\rho)(\delta-1)\eta)\zeta - (d+\alpha+\gamma)\big)\,\beta'(\rho) + \eta\zeta\beta(\rho)(\delta-1)\big)S^*}{(d+\alpha+\gamma)(\zeta+d+\alpha+\gamma)} \quad \text{S31}$$

#### 2 Stability analysis of singular strategies

##### *2.1 Model A*

The fitness gradient for model A is,

$$\mathcal{F}(\rho) = \frac{-S^*\left((-1+\eta(\delta-1)(\rho-1)\sigma)\frac{d\beta(\rho)}{d\rho} + \beta(\rho)\eta\sigma(\delta-1)\right)}{d+\alpha+\gamma} \quad \text{S32}$$

and the density of susceptibles at steady state is,

$$S^* = \frac{-(d+\alpha+\gamma)}{(-1+\eta(\delta-1)(-1+\rho)\sigma)\beta(\rho)} \quad \text{S33}$$

with derivative

$$\frac{dS}{d\rho} = \frac{-(d+\alpha+\gamma)\eta(\delta-1)\sigma}{(-1+\eta(\delta-1)(-1+\rho)\sigma)^2\beta(\rho)} - \frac{-(d+\alpha+\gamma)\frac{d\beta(\rho)}{d\rho}}{(-1+\eta(\delta-1)(-1+\rho)\sigma)\beta(\rho)^2} \quad \text{S34}$$

At the singular strategy the fitness gradient is zero, and hence we know,

$$\left.\frac{d\beta(\rho)}{d\rho}\right|_{\rho=\rho^*} = \frac{-\sigma\eta(\delta-1)\beta(\rho^*)}{-1+\eta(-1+\rho^*)(\delta-1)\sigma} \quad \text{S35}$$

Hence, substituting in expression S35 into S34 gives,

$$\left.\frac{dS}{d\rho}\right|_{\rho=\rho^*} = \frac{(d+\alpha+\gamma)\eta(\delta-1)\sigma}{(-1+\eta(\delta-1)(-1+\rho^*)\sigma)^2\beta(\rho^*)} - \frac{(d+\alpha+\gamma)\sigma\eta(\delta-1)}{(-1+\eta(\delta-1)(-1+\rho^*)\sigma)^2\beta(\rho^*)} = 0 \quad \text{S36}$$

The convergence stability of a singular strategy $\rho^*$ is determined by the mutual invaisability $M$, and the evolutionary stability $E$, where $M = \left.\frac{\partial^2 r(\rho_m,\rho)}{\partial\rho\partial\rho_m}\right|_{\rho=\rho_m=\rho^*}$ and $E = \left.\frac{\partial^2 r(\rho_m,\rho)}{\partial\rho_m^2}\right|_{\rho=\rho_m=\rho^*}$. A singular strategy is convergence stable when $E+M<0$ and evolutionarily stable when $E<0$. Here,

$$M = \left.\frac{\partial^2 r(\rho_m,\rho)}{\partial\rho\partial\rho_m}\right|_{\rho=\rho_m=\rho^*} = \frac{-\left((-1+\eta(\delta-1)(\rho^*-1)\sigma)\frac{d\beta(\rho)}{d\rho}|_{\rho^*} + \beta(\rho^*)\eta\sigma(\delta-1)\right)}{d+\alpha+\gamma}\left.\frac{dS}{d\rho}\right|_{\rho=\rho^*} = 0 \quad \text{S37}$$

Hence, any singular strategy is convergence stable if it is evolutionarily stable.

#### 2.2 Model B

The fitness gradient in Model B is,

$$\mathcal{F}(\rho) = \frac{1}{(d+\alpha+\gamma)(\zeta(1-\rho)+d+\alpha+\gamma)^2}\Big(\big((\zeta(1-\rho)+d+\alpha+\gamma)(-(1+(\delta-1)\eta)(\rho-1)\zeta+d+\alpha+\gamma)\beta'(\rho) - \eta\zeta\beta(\rho)(\delta-1)(d+\alpha+\gamma)\big)S^*\Big) \quad \text{S38}$$

And the density of susceptibles is,

$$S = \frac{(\zeta(1-\rho)+d+\alpha+\gamma)(d+\alpha+\gamma)}{\beta(\rho)(-1(1+(\delta-1)\eta)(-1+\rho)\zeta+d+\alpha+\gamma)} \quad \text{S39}$$

with derivative

$$\frac{dS}{d\rho} = \frac{-1}{\beta(\rho)^2(-1(-1+\rho)(1+(\delta-1)\eta)\zeta+d+\alpha+\gamma)^2}\left(((\zeta(1-\rho)+d+\alpha+\gamma)(-1(-1+\rho)(1+(\delta-1)\eta)\zeta+d+\alpha+\gamma)\frac{d\beta(\rho)}{d\rho} - \eta\zeta\beta(\rho)(d+\alpha+\gamma)(\delta-1))(d+\alpha+\gamma)\right) \quad \text{S40}$$

At the singular strategy $\rho^*$, the fitness gradient is 0, and hence,

$$\left.\frac{d\beta(\rho)}{d\rho}\right|_{\rho=\rho^*} = \frac{\eta\zeta\beta(\rho^*)(\delta-1)(d+\alpha+\gamma)}{(\zeta(1-\rho^*)+d+\alpha+\gamma)(-1(1+(\delta-1)\eta)(-1+\rho^*)\zeta+d+\alpha+\gamma)} \quad \text{S41}$$

Substituting S41 into S40, we obtain

$$\left.\frac{dS}{d\rho}\right|_{\rho=\rho^*} = \frac{-1}{\beta(\rho^*)^2(-1(-1+\rho^*)(1+(\delta-1)\eta)\zeta+d+\alpha+\gamma)^2}\Big(((\zeta(1-\rho^*)+d+\alpha+\gamma)(-1(-1+\rho^*)(1+(\delta-1)\eta)\zeta+d+\alpha+\gamma)\frac{\eta\zeta\beta(\rho^*)(\delta-1)(d+\alpha+\gamma)}{(\zeta(1-\rho^*)+d+\alpha+\gamma)(-1(1+(\delta-1)\eta)(-1+\rho^*)\zeta+d+\alpha+\gamma)} - \eta\zeta\beta(\rho^*)(d+\alpha+\gamma)(\delta-1))(d+\alpha+\gamma)\Big) \quad \text{S42}$$

$$\left.\frac{dS}{d\rho}\right|_{\rho=\rho^*} = \frac{-1}{\beta(\rho^*)^2(-1(-1+\rho^*)(1+(\delta-1)\eta)\zeta+d+\alpha+\gamma)^2}(\eta\zeta\beta(\rho^*)(\delta-1)(d+\alpha+\gamma) - \eta\zeta\beta(\rho^*)(d+\alpha+\gamma)(\delta-1))(d+\alpha+\gamma)) = 0 \quad \text{S43}$$

Here,

$$M = \left.\frac{\partial^2 r(\rho_m, \rho)}{\partial\rho\partial\rho_m}\right|_{\rho=\rho_m=\rho^*} = \frac{1}{(d+\alpha+\gamma)(\zeta(1-\rho)+d+\alpha+\gamma)^2}\Big(\big(((\zeta(1-\rho)+d+\alpha+\gamma)(-(1+(\delta-1)\eta)(\rho-1)\zeta+d+\alpha+\gamma)\beta'(\rho) - \eta\zeta\beta(\rho)(\delta-1)(d+\alpha+\gamma)\big)\Big)\left.\frac{dS}{d\rho}\right|_{\rho=\rho^*} = 0 \tag{S44}$$

Hence any evolutionarily stable singular strategy is also convergent stable.

#### 2.3 Model C

The fitness gradient in Model C is,

$$\mathcal{F}(\rho) = \frac{\big((-1+(-1+\rho)(\delta-1)\eta)\zeta-(d+\alpha+\gamma)\big)\beta'(\rho)+\eta\zeta\beta(\rho)(\delta-1)\big)S^*}{(d+\alpha+\gamma)(\zeta+d+\alpha+\gamma)} \tag{S45}$$

And the density of susceptibles is,

$$S = \frac{(\zeta+d+\alpha+\gamma)(d+\alpha+\gamma)}{\big((1-(\delta-1)(-1+\rho)\eta)\zeta+d+\alpha+\gamma\big)\beta(\rho)} \tag{S46}$$

With derivative

$$\frac{dS}{d\rho} = \frac{\Big(\big((1-(\delta-1)(-1+\rho)\eta)\zeta+d+\alpha+\gamma\big)\beta'(\rho)-\eta\zeta\beta(\rho)(\delta-1)\Big)(\zeta+d+\alpha+\gamma)(d+\alpha+\gamma)}{\big((1-(\delta-1)(-1+\rho)\eta)\zeta+d+\alpha+\gamma\big)^2\beta(\rho)^2} \tag{S45}$$

At the singular strategy $\rho^*$, the fitness gradient is 0, and hence,

$$\left.\frac{d\beta(\rho)}{d\rho}\right|_{\rho=\rho^*} = \frac{\eta\zeta\beta(\rho^*)(\delta-1)}{(1-(-1+\rho^*)(\delta-1)\eta)\zeta+d+\alpha+\gamma} \tag{S46}$$

Substituting S46 into S45, we obtain

$$\left.\frac{dS}{d\rho}\right|_{\rho=\rho^*} = \frac{\left(\big((1-(\delta-1)(-1+\rho^*)\eta)\zeta+d+\alpha+\gamma\big)\frac{\eta\zeta\beta(\rho^*)(\delta-1)}{(1-(-1+\rho^*)(\delta-1)\eta)\zeta+d+\alpha+\gamma}-\eta\zeta\beta(\rho^*)(\delta-1)\right)(\zeta+d+\alpha+\gamma)(d+\alpha+\gamma)}{\big((1-(\delta-1)(-1+\rho^*)\eta)\zeta+d+\alpha+\gamma\big)^2\beta(\rho^*)^2} \tag{S47}$$

$$\left.\frac{dS}{d\rho}\right|_{\rho=\rho^*} = \frac{\big(\eta\zeta\beta(\rho^*)(\delta-1)-\eta\zeta\beta(\rho^*)(\delta-1)\big)(\zeta+d+\alpha+\gamma)(d+\alpha+\gamma)}{\big((1-(\delta-1)(-1+\rho^*)\eta)\zeta+d+\alpha+\gamma\big)^2\beta(\rho^*)^2} = 0 \tag{S48}$$

Here

$$M = \frac{\partial^2 r(\rho_m, \rho)}{\partial\rho\partial\rho_m}\bigg|_{\rho=\rho^*}$$
$$= \frac{\big((-1 + (-1 + \rho)(\delta - 1)\eta)\zeta - (d + \alpha + \gamma)\big)\,\beta'(\rho) + \eta\zeta\beta(\rho)(\delta - 1)\big)}{(d + \alpha + \gamma)(\zeta + d + \alpha + \gamma)}\frac{dS}{d\rho}\bigg|_{\rho=\rho^*}$$
$$= 0 \quad \text{S49}$$

Hence any evolutionarily stable singular strategy is also convergence stable.

### 3 Selection for detection avoidance

#### 3.1 Model A

##### 3.1.1 Perfect compliance and quarantining

The fitness gradient for model A when $\delta = 0, \eta = 1$ is,

$$\mathcal{F}(\rho) = \frac{-S^*\left((-1 - (\rho - 1)\sigma)\frac{d\beta(\rho)}{d\rho} - \beta(\rho)\sigma\right)}{d + \alpha + \gamma} \quad \text{S50}$$

There is selection for detection avoidance when $\mathcal{F}(0) > 0$:

$$\mathcal{F}(0) = \frac{-S^*\left((-1 - (0 - 1)\sigma)\frac{d\beta(\rho)}{d\rho}\bigg|_{\rho=0} - \beta(0)\sigma\right)}{d + \alpha + \gamma}$$
$$= \frac{-S^*}{d + \alpha + \gamma}\left((\sigma - 1)\frac{d\beta(\rho)}{d\rho}\bigg|_{\rho=0} - \beta(0)\sigma\right) > 0 \quad \text{S51}$$

Hence,

$$(\sigma - 1)\frac{d\beta(\rho)}{d\rho}\bigg|_{\rho=0} - \beta(0)\sigma < 0 \quad \text{S52}$$

We can rearrange S52 in terms of $\sigma$ to find the minimum testing probability needed for detection avoidance to evolve, which is:

$$\sigma_{min} > \frac{\frac{d\beta(\rho)}{d\rho}\Big|_{\rho=0}}{\frac{d\beta(\rho)}{d\rho}\Big|_{\rho=0} - \beta(0)} \quad \text{S53}$$

##### 3.1.2 Imperfect quarantining

The fitness gradient for model A is,

$$\mathcal{F}(\rho) = \frac{-S^*\left((-1+\eta(\delta-1)(\rho-1)\sigma)\frac{d\beta(\rho)}{d\rho} + \beta(\rho)\eta\sigma(\delta-1)\right)}{d+\alpha+\gamma} \tag{S54}$$

As above, there is selection for detection avoidance when $\mathcal{F}(0) > 0$:

$$\begin{aligned}\mathcal{F}(0) &= \frac{-S^*\left((-1+\eta(\delta-1)(0-1)\sigma)\frac{d\beta(\rho)}{d\rho}\Big|_{\rho=0} + \beta(0)\eta\sigma(\delta-1)\right)}{d+\alpha+\gamma} \\ &= \frac{-S^*}{d+\alpha+\gamma}\left((-1-\eta(\delta-1)\sigma)\frac{d\beta(\rho)}{d\rho}\Big|_{\rho=0} - \beta(0)\eta(\delta-1)\sigma\right) \\ &> 0\end{aligned} \tag{S55}$$

Hence,

$$(-1-\eta(\delta-1)\sigma)\frac{d\beta(\rho)}{d\rho}\Big|_{\rho=0} - \beta(0)\eta(\delta-1)\sigma < 0 \tag{S56}$$

We can rearrange S56 in terms of $\sigma$ to find the minimum testing probability needed for detection avoidance to evolve:

$$\sigma_{min} > \frac{-\frac{d\beta(\rho)}{d\rho}\Big|_{\rho=0}}{\eta(\delta-1)\left(\frac{d\beta(\rho)}{d\rho}\Big|_{\rho=0} - \beta(0)\right)} \tag{S57}$$

#### *3.2 Model B*

##### *3.2.1 Perfect compliance and quarantining*

The fitness gradient for Model B when there is perfect compliance and quarantining is,

$$\mathcal{F}(\rho) = \frac{d\beta(\rho)}{d\rho}\left(\frac{\zeta(1-\rho)+d+\alpha+\gamma}{\beta(\rho)}\right) + \zeta \tag{S58}$$

There is selection for detection avoidance when $\mathcal{F}(0) > 0$:

$$\mathcal{F}(0) = \frac{d\beta(\rho)}{d\rho}\Big|_{\rho=0}\left(\frac{\zeta+d+\alpha+\gamma}{\beta(0)}\right) + \zeta > 0 \tag{S59}$$

We can rearrange this to find the minimum testing rate needed for detection avoidance to evolve:

$$\zeta > \frac{-\left.\frac{d\beta(\rho)}{d\rho}\right|_{\rho=0}(d+\alpha+\gamma)}{\left.\frac{d\beta(\rho)}{d\rho}\right|_{\rho=0}+\beta(0)} \quad \text{S60}$$

*3.2.2 Imperfect quarantining*

The fitness function for the general case is

$$\mathcal{F}(\rho) = \frac{1}{(d+\alpha+\gamma)(\zeta(1-\rho)+d+\alpha+\gamma)^2}\left(\left((\zeta(1-\rho)+d+\alpha+\gamma)(-(1+(\delta-1)\eta)(\rho-1)\zeta+d+\alpha+\gamma)\frac{d\beta(\rho)}{d\rho} - \eta\zeta\beta(\rho)(\delta-1)(d+\alpha+\gamma)\right)S^*\right) \quad \text{S61}$$

Here we will write $\tilde{d} = d + \alpha + \gamma$ for brevity, hence

$$\mathcal{F}(\rho) = \frac{1}{\tilde{d}\left(\zeta(1-\rho)+\tilde{d}\right)^2}\left(\left(\left(\zeta(1-\rho)+\tilde{d}\right)\left(-(1+(\delta-1)\eta)(\rho-1)\zeta+\tilde{d}\right)\frac{d\beta(\rho)}{d\rho} - \eta\zeta\beta(\rho)(\delta-1)\tilde{d}\right)S^*\right) \quad \text{S62}$$

Selection favours detection avoidance when $\mathcal{F}(0) > 0$:

$$\mathcal{F}(0) = \frac{1}{\tilde{d}\left(\zeta+\tilde{d}\right)^2}\left(\left(\left(\zeta+\tilde{d}\right)\left(-(1+(\delta-1)\eta)(-1)\zeta+\tilde{d}\right)\left.\frac{d\beta(\rho)}{d\rho}\right|_{\rho=0} - \eta\zeta\beta(0)(\delta-1)\tilde{d}\right)S^*\right) > 0 \quad \text{S63}$$

Hence,

$$\left(\zeta+\tilde{d}\right)\left(-(1+(\delta-1)\eta)(-1)\zeta+\tilde{d}\right)\left.\frac{d\beta(\rho)}{d\rho}\right|_{\rho=0} - \eta\zeta\beta(0)(\delta-1)\tilde{d} > 0 \quad \text{S64}$$

This can be rearranged to obtain a quadratic in $\zeta$, which can be solved to determine upper and lower bounds on testing rates for detection avoidance to evolve.

#### 3.3 Model C

##### 3.3.1 Perfect compliance and quarantining

The fitness gradient for Model C when there is perfect compliance and quarantining is,

$$\mathcal{F}(\rho) = \frac{\big(\rho\zeta + (d+\alpha+\gamma)\big)\,\beta'(\rho) + \zeta\beta(\rho)\big)S^*}{(d+\alpha+\gamma)(\zeta+d+\alpha+\gamma)} \quad \text{S65}$$

There is selection for detection avoidance when $\mathcal{F}(0) > 0$, hence

$$\mathcal{F}(0) = (d+\alpha+\gamma)\left.\frac{d\beta(\rho)}{d\rho}\right|_{\rho=0} + \zeta\beta(0) > 0 \quad \text{S66}$$

We can rearrange this to find the minimum testing rate needed for detection avoidance to evolve:

$$\zeta_{min} > \frac{-(d+\alpha+\gamma)\left.\frac{d\beta(\rho)}{d\rho}\right|_{\rho=0}}{\beta(0)} \quad \text{S67}$$

##### 3.3.2 Imperfect quarantining

The fitness function for the general case is

$$\mathcal{F}(\rho) = \frac{\big((-1+(-1+\rho)(\delta-1)\eta)\zeta - (d+\alpha+\gamma)\big)\,\beta'(\rho) + \eta\zeta\beta(\rho)(\delta-1)\big)S^*}{(d+\alpha+\gamma)(\zeta+d+\alpha+\gamma)} \quad \text{S68}$$

Here we will write $\tilde{d} = d+\alpha+\gamma$ for brevity, hence

$$\mathcal{F}(\rho) = \frac{\Big((-1+(-1+\rho)(\delta-1)\eta)\zeta - \tilde{d}\big)\,\beta'(\rho) + \eta\zeta\beta(\rho)(\delta-1)\Big)S^*}{\tilde{d}(\zeta+\tilde{d})} \quad \text{S69}$$

Selection favours detection avoidance when $\mathcal{F}(0) > 0$:

$$\mathcal{F}(0) = \frac{\left((-1-(\delta-1)\eta)\zeta - \tilde{d}\right)\left.\frac{d\beta(\rho)}{d\rho}\right|_{\rho=0} + \eta\zeta\beta(0)(\delta-1)\Bigg)S^*}{\tilde{d}(\zeta+\tilde{d})} > 0 \quad \text{S70}$$

Hence,

$$\zeta_{min} > \frac{\tilde{d}\left.\frac{d\beta(\rho)}{d\rho}\right|_{\rho=0}}{\left.\frac{d\beta(\rho)}{d\rho}\right|_{\rho=0}(-1-\delta\eta+\eta) + \eta\beta(0)(\delta-1)} \quad \text{S71}$$

### 4 Supplementary figures

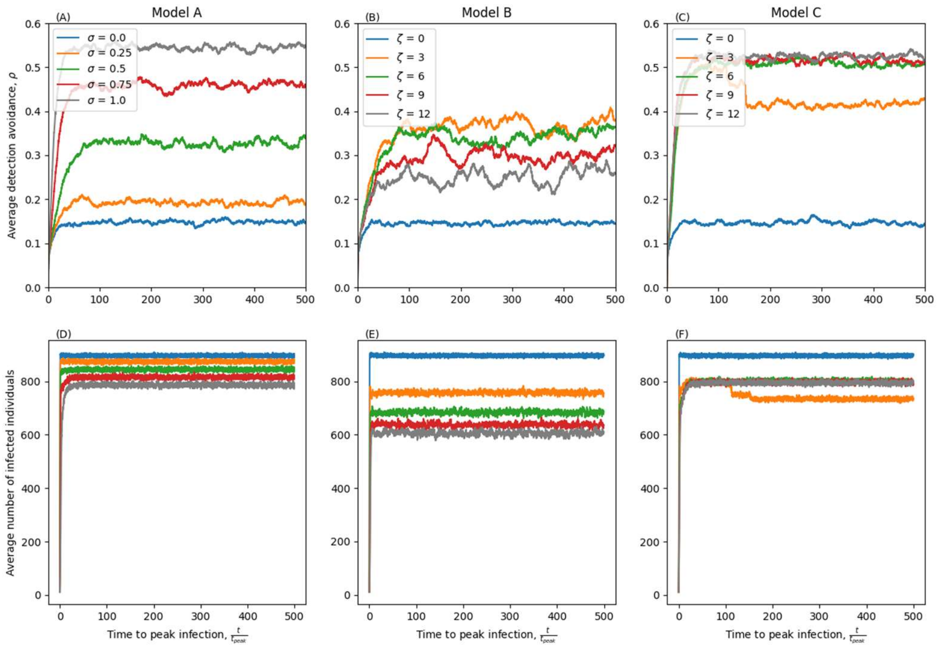


*Figure S1: Stochastic simulations of dynamics for Models A-C (A-C). The first row of figures show the average detection avoidance. The second row of figures show the average number of infected individuals. Parameters as in Table 1 except $\delta$=0.1, $\eta$=0.90, $\beta$=0.01. Discard threshold is 100 infected individuals.*

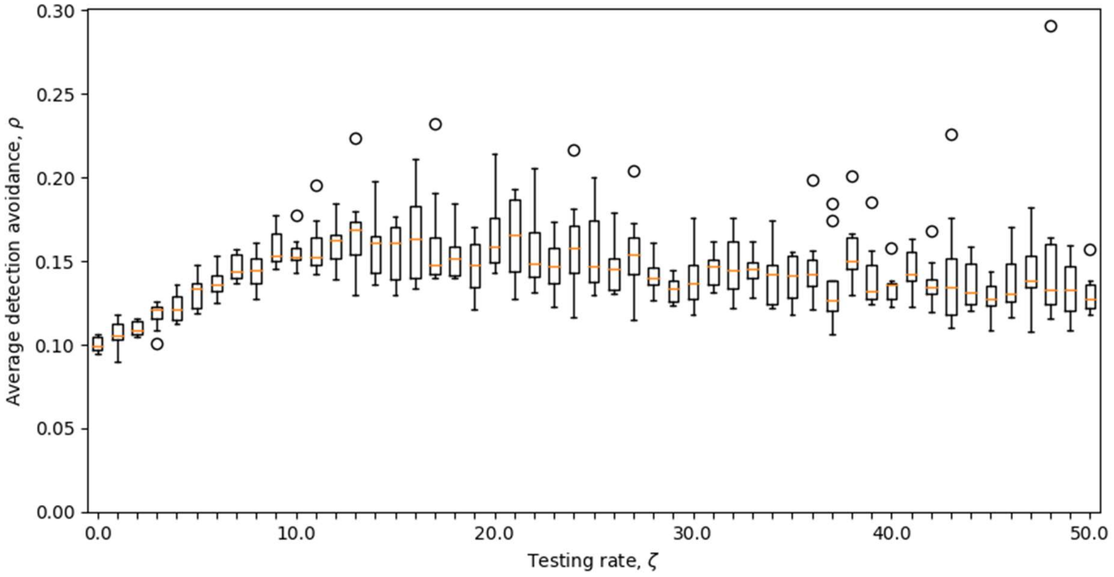


*Figure S2: Box plot of average detection avoidance $\rho$ at peak number of infected individuals for Model B. Parameters as in Table 1 except $\delta = 0.1, \eta = 0.9, \beta = 0.00001$.*
